# Attitudes and beliefs about family and domestic violence in Australian faith-based communities: A qualitative study

**DOI:** 10.1101/2020.10.29.20222703

**Authors:** Mandy Truong, Mienah Sharif, Anna Olsen, Dave Pasalich, Bianca Calabria, Naomi Priest

## Abstract

Family and domestic violence (FDV) is a major public health and social issue that is associated with a range of physical, mental and behavioural health outcomes. Religion and faith are powerful and influential in shaping the lives of many individuals and societies, in addition to the social practices, norms and structures that are significant in understanding and responding to FDV. This qualitative study aims to deepen understanding of the influence of religious beliefs and values on attitudes and beliefs of FDV among culturally diverse faith communities in Australia. Interviews and focus groups were conducted with 64 participants from a diverse range of cultural and religious backgrounds which included faith leaders, community members and FDV sector workers. Six main themes were identified describing attitudes, beliefs and knowledge about FDV: 1) Faith and religion do not condone violence; 2) Awareness of FDV is increasing, yet remains often poorly understood; 3) FDV is still a taboo topic; 4) Denial and defensiveness about FDV persist; 5) Patience, endurance and forgiveness is often prioritised over safety; 6) Gender roles and norms founded on religious beliefs and interpretations underpin many FDV understandings and responses. These findings demonstrate the tensions between expressions of faith and attitudes to women and FDV. Further exploration of these issues within specific faith communities, as well as how to support and engage with these communities in increasing understandings of FDV and developing effective responses, is needed in the Australian context.

## Introduction

Family and domestic violence (FDV) is a major public health issue with immediate and long term impacts on women’s physical, mental, behavioural and reproductive health (World Health Organization, 2012). Children exposed to FDV are at increased risk of learning and developmental difficulties, and mental health and behavioural issues in childhood and throughout later life (UNICEF, 2006). Wider societal impacts of FDV are also substantial, including homelessness for women and children (DiNicola, Liyanarachchi, & Plummer, 2018), and reduced job satisfaction and work productivity (Banyard, Potter, & Turner, 2011; Breiding, Chen, & Black, 2014). Thirty percent of women globally have reported physical and/or sexual violence by their partners at least once in their lifetime (World Health Organization, 2013). One in four Australian women has experienced physical or sexual violence by a current of former partner since the age of fifteen years and one in four Australian women has experienced emotional abuse by a current or former partner (Australian Bureau of Statistics, 2017a). Almost ten Australian women a day are hospitalized for assault injuries perpetrated by a spouse or domestic partner, and on average one Australian woman a week is murdered by her current or former partner (Australian Institute of Health and Welfare, 2019; Bryant & Bricknell, 2017; Domestic Violence Death Review Team, 2018).

Throughout the world and across settings, FDV is defined and referred to in various ways, with domestic violence and intimate partner violence often used interchangeably to refer to violence between people who have, or have had, an intimate relationship; family violence is a broader term that includes violence between intimate partners and between family members (World Health Organization, 2012). In Australia, the term family violence is used to recognize the wide range of marital and kinship relationships in which violence may occur for Indigenous people (Department of Social Services, 2014). This also applies to some ethnic minoritized and migrant groups. Consistent with this approach the term family and domestic violence is used throughout this paper.

Australia is one of the most ethnically and culturally diverse countries globally and with a greater proportion of the population born overseas than the United States, Canada and New Zealand (Australian Bureau of Statistics, 2020), however, empirical studies on family and domestic violence among women from migrant and refugee backgrounds is limited. This includes a lack of comprehensive, population-wide data on the prevalence and impacts of violence against women from migrant and refugee backgrounds. Existing community-based studies suggest high prevalence rates and specific issues of complexity related to the use of temporary visa status as a means of violence, communication and access to culturally appropriate information and services, and religious abuse (Australian Institute of Health and Welfare, 2018; Vaughan et al., 2016). Although existing community-based studies on migrant and refugee women’s experiences of FDV provide valuable insights, the influence of religion and faith practices and beliefs on FDV has received little research attention in Australia.

More than two thirds of all Australians report affiliation with a faith or religion. Christianity is the largest faith affiliation reported (52%), followed by Islam (2.6%) and Buddhism (2.4%). Australia’s religious diversity is increasing, with the proportion of Australians reporting a religious affiliation other than Christian increasing from 5.6% in 2006 to 8.2% in 2016. This was spread across most non-Christian religions, with Islam (1.7% to 2.6%) and Hinduism (0.7% to 1.9%) showing the highest changes (Australian Bureau of Statistics, 2017b). In 2019, 29.7% of the Australian population were born overseas, a higher proportion than the United States, Canada, or New Zealand (Australian Bureau of Statistics, 2020). Religion, culture and experiences of migration intersect in complex ways, and disentangling religious and cultural norms is difficult (Le Roux, 2015; Vaughan et al., 2020). Recognition of the interconnectedness of religion and culture should not however be seen as only applicable to migrant communities (Priest, 2018; Vaughan et al., 2020). The role of cultural norms in FDV and the need for wider cultural change to end this violence needs urgent attention within the whole of Australian society, including in more established faith communities (Priest, 2018; Vaughan et al., 2020).

### Religion and Family and Domestic Violence

Religion, faith and faith-based communities play central roles in the lives of many people and faith leaders provide moral, spiritual and social support for their communities, including support for those experiencing FDV (IMA World Health & Sojourners, 2018; Our Watch, ANROWS, & VicHealth, 2015; Vaughan et al., 2020). Faith leaders and faith-communities have potential to promote norms and relationships founded on respect and equality that protect against FDV. Faith leaders can be highly influential in shaping attitudes and behaviours related to FDV due to the trust and authority they hold within their communities (Australian Muslim Women’s Centre for Human Rights, 2011; CHALLENGE Family Violence project, 2015; Jewish Taskforce Against Family Violence, 2011; State of Victoria, 2016). They may also promote norms and relationships that drive or condone violence. Outstanding knowledge gaps regarding the causes and drivers of FDV among religious and faith-based communities, and about effective strategies to respond to and prevent such violence have been recognized within the Australian context (Australian Bureau of Statistics, 2013; Bartels, 2010; Department of Social Services, 2016; Vaughan et al., 2020a).

In many contexts and communities throughout the world, religion and faith communities inform gender norms and beliefs in both public and private/domestic spaces, although are often contested (Adjei & Mpiani, 2020; McMorris & Glass, 2018). Despite a considerable body of work exploring the links between culture and FDV, fewer studies have examined the role of religion specifically in relation to FDV (Adjei & Mpiani, 2020; Priest, 2018). Work that does exist in Western countries is almost exclusively from the United States and among Christian groups, with far less in other contexts (Aune & Barnes, 2018; Le Roux, 2015). Many faith-based groups are keen to engage in work to prevent and address FDV, and there is an outstanding need to better understand the relevant cultural beliefs and practices across different religious groups and its impact on prevention and responses to FDV (Le Roux, 2015).

Existing empirical evidence highlights the complex influence of religious beliefs and values on attitudes, experiences and responses to FDV. Some studies have suggested that men who attend church regularly are less likely to perpetrate domestic violence (Fergusson, Horwood, Kershaw, & Shannon, 1986; Wilcox, 2004), although these studies are dated and do not capture non-physical forms of abuse (Priest, 2018). Studies have found no difference in rates of abuse between those attending and not attending church (Institute for Family Studies & Wheatley Institution, 2019; Wang, Horne, Levitt, & Klesges, 2009).

Among some scholars, faith and religion is viewed as a protective factor against FDV through promotion of traditional family values and limiting of alcohol and other substance use which are risk factors for FDV (Adjei & Mpiani, 2020; Takyi & Lamptey, 2020). Moreover, faith and religious beliefs have been identified as important coping mechanisms for women experiencing FDV. Prayer and faith texts are key sources of comfort and support for many women of faith (Beaulaurier, Seff, Newman, & Dunlop, 2007; Berry & York, 2011; McIntosh & Rosselli, 2012; Westenberg, 2017) and spirituality is a potential buffer against harms associated with FDV (de la Rosa, Barnett-Queen, Messick, & Gurrola, 2016; Drumm et al., 2014). Higher levels of spirituality and greater religious involvement can be associated with fewer depression symptoms and better mental health, including among survivors of FDV (Koenig, 2012; Watlington & Murphy, 2006). Faith leaders and communities are also sources of support for women of faith backgrounds experiencing violence (Gillum, Sullivan, & Bybee, 2006; Holmes, 2012; Vaughan et al., 2016).

In contrast, other studies identify factors within religion and faith-communities that support drivers of FDV. These include an emphasis on patriarchy and conceptions of gender and appropriate behaviour for men and women in intimate relationships together with institutional norms that legitimise male dominance (Adjei & Mpiani, 2020; Eidhamar, 2018; Hindelang, 2000; Ringel & Park, 2008; Westenberg, 2017). Religiosity can legitimize and predict acceptance of FDV through beliefs and teachings that emphasise male authority and female submission. Male perpetrators of violence against women have been found to justify their behaviour based on patriarchal religious ideologies and interpretations of sacred texts (Le Roux & Bowers-Du Toit, 2017; Lock, 2018; Ringel & Park, 2008). Women in faith communities who experience violence often use religious language and symbolism to explain and tolerate violence, and remain in abusive relationships longer than other women (Hosseini-Sedehi, 2016; Knickmeyer, Levitt, & Horne, 2010; Popescu et al., 2009; Westenberg, 2017).

At the institutional level, several factors have been identified which may enable the perpetration of FDV including: denial of FDV, or minimization of its severity, within the faith; silencing of women who disclose experiences of violence; and inappropriate responses to disclosure of abuse such as providing marriage counselling (IMA World Health & Sojourners, 2018; Le Roux, 2015; Sojourners & IMA World Health, June 2014). For example, women who disclose abusive relationships can be counselled by religious leaders to prioritise a ‘faith first’ approach, such as prayer and church attendance, over their own safety (Lock, 2018). The extent to which religious institutions can contribute to addressing FDV and changing unequal gender relations while excluding women within their own leadership structures has been raised as a key outstanding issue (Holmes, 2012; Vaughan et al., 2020; Westenberg, 2017) Finally, a lack of engagement with faith communities by secular FDV care providers and an unwillingness or poor skills among secular services in supporting religious survivors of abuse also impact the ability of faith leaders and faith communities to prevent and respond to FDV (IMA World Health & Sojourners, 2018; Le Roux, 2015; Vaughan et al., 2020).

Much of the existing research on family violence is drawn from contexts outside of Australia where population and migration demographics and wider cultural, religious and gender norms are markedly different. These past studies are dated and there is a lack of research conducted in recent years. In this light, there is a pressing need for contemporary evidence regarding religion and FDV in Australia. This study aims to contribute to empirical gaps in the literature by exploring attitudes, beliefs and knowledge about FDV in culturally diverse faith-based communities in Australia in an in-depth qualitative study design.

## Method

### Research framework and approach

Qualitative methods, underpinned by a social constructionist approach, were used in this study to explore the perspectives of participants (Bourgeault, Dingwall, & De Vries, 2010; Liamputton & Ezzy, 2005). This approach seeks to understand how people make sense of their lives by studying the lived experience of a variety of people under different circumstances (Yin, 2015). FDV sector workers, faith leaders, and community members from a range of faiths and religions and racial/ethnic backgrounds were sought to gain multiple understandings of FDV.

### Ethics, informed consent and safety protocol

Ethics approval was obtained from the Australian National University’s Human Research Ethics Committee (HREC; Protocol #2018/316). In consideration of religious and cultural sensitivities, the researchers consulted with FDV sector stakeholders and other contacts. They were given the opportunity to provide feedback on the interview and focus group questions and study procedures to ensure they were appropriate and acceptable. For example, use of appropriate terminology such as ‘respectful relationships’ and ‘family safety’ were used rather than academic terminology at times to facilitate discussion related to FDV.

A safety protocol was developed which outlined strategies to minimise the risk of participants becoming distressed during their participation in our study. This included emphasising to each participant that their involvement was entirely voluntary and that they were not required to answer any questions they did not wish to. Researchers endeavoured to conduct all interviews and focus groups in a supportive and non-judgemental manner. Potential risks for researchers were also identified. Researchers conducting data collection had regular debriefs with supervisors and access to a confidential counselling service.

### Recruitment and data collection procedures

Convenience and purposive sampling strategies were used to recruit participants to obtain a range of views from: individuals working in the FDV sector, faith leaders and faith community members. Firstly, existing relationships and networks were used to identify relevant people and organisations to contact. Second, purposeful sampling was used to target particular faiths were not representation in the sample. Stakeholders and leaders from FDV organisations, related agencies (e.g. community health), and faith communities were contacted and asked to identify potential participants to invite to participate in the study. All participants resided in one of four Australian states (Victoria, New South Wales, Queensland and South Australia).

Participants were provided with a plain language statement describing the project and had opportunity to ask questions about the project and their involvement. Plain language statements included a list of support services that participants could contact should they require support and assistance. Written or verbal informed consent was obtained from all participants.

Interviews and focus groups were conducted from August to November 2018. On average, interviews (face-to-face or via phone) and focus groups (face-to-face; up to nine participants in each focus group) were between 30 and 60 minutes. Focus groups were usually arranged with the assistance of a community worker or advocate that was known to the community members and who was often present during the group discussion. Interviews and focus groups were audio-recorded and transcribed with consent from participants and field notes were made by the researcher facilitating the data collection during and straight after the interview or focus group.

### Interview topics

Semi-structured interview guides were developed by the researchers in order to understand participants’ perceptions and experiences of FDV in faith-based communities. Faith leaders were asked about their awareness and perception of the prevalence of FDV in their communities, and what key issues were faced by their community in relation to FDV. Leaders were also asked about any programs or training they were involved in that addressed FDV and what gaps needed to be addressed. Interviews and focus group discussions with faith community members focussed on their perspectives of the awareness and prevalence of family violence in their communities, and how much of an issue they felt it was among their community. Faith community member participants were also asked about the role of their faith leader within their community and their perceptions of faith leaders’ capacity to address FDV. Participants were not asked to disclose personal experiences of family violence. Interviews with FDV sector workers explored their experiences working with faith-based communities in FDV in terms of the types of communities they had worked with and what kinds of attitudes, beliefs and experiences they had encountered. FDV sector worker participants were also asked their perceptions of how faith leaders and communities responded to FDV and what issues they thought were important for faith communities.

### Participants

Interviews were conducted with eleven faith leaders who self-identified as being from the following faiths: Catholicism, Christianity (Catholic, Evangelical and Anglican), Islam, Buddhism and Judaism. Their self-identified cultural backgrounds included: Sudanese, Sri Lankan, Chinese, Afghan and Jewish. All, except one, were individually interviewed (face-to-face or via phone). One faith leader was part of a focus group with community members. Three of the eleven faith leaders were women. Some of the faith leaders consulted had been involved in FDV related work within their communities, such as conducting workshops to raise awareness of FDV.

Interviews and focus groups were conducted with a total of 47 female and male faith community members from the following self-identified faiths: Christianity, Islam, Buddhism, Hindu and Judaism. Their self-identified cultural backgrounds included: Indian, Pakistani, Jewish, Lebanese, Afghan and Vietnamese. Some community members had experience working in the FDV sector (e.g. as activists, community workers.)

Interviews were conducted with seven FDV sector workers (four male and three female) from a range of organisations including: FDV advocacy organisations, tertiary religious studies units, and community social services. These FDV sector worker participants worked with leaders and communities from the following faiths communities: Catholic, Christian, Buddhist, Muslim, Hindu and Sikh. Some also had a faith-background themselves and were working in FDV either within their faith community or with other faith communities. One FDV sector worker had been a faith leader previously.

### Participation and recruitment issues

Overall, there was a willingness from staff working at social service agencies and advocacy groups to engage in the study, and a strong view articulated that more work was needed to understand the issues related to FDV faced by faith communities in Australia. In contrast, recruitment of faith leader and community member participants was more challenging. Some faith leaders and community groups were wary of participating in the project, providing feedback that they felt they were already over-consulted without any resulting practical outcomes for their communities. Several Islamic-faith groups reported feeling that their communities were being targeted using a deficit-framework and were concerned that it would contribute to further negatively stereotyping their faith and cultural group. In addition, a number of potential participants expressed that explicitly discussing the topic of FDV is discouraged, or even stigmatized, in many faith-based and ethnic communities so they were reluctant to participate in the consultations. As a result, several of the community member consultations were conducted via individual interviews rather than focus groups as some people felt more comfortable discussing without others present.

### Data analysis

Inductive and deductive thematic analysis was undertaken using three phases: preparation, organization and reporting (Elo & Kyngäs, 2008; Hsieh & Shannon, 2005). Qualitative analysis software NVIVO was used for data management and coding (QSR International, 2015). The first author read and openly coded all transcripts and categories were generated. After the process of open coding, related categories were grouped under higher order ‘nodes’ (i.e. categories) to develop a coding nodes structure. Nodes were analysed into themes. The first, second and last author discussed emerging patterns, nodes and themes. All authors were involved in discussion and finalising of the themes. In addition, representatives from the Australian Government Department of Social Services (funders of the project) were also involved in discussion of emerging themes during data analysis and also reviewed and provided feedback on the final themes.

### Researcher context

An implication of using a social constructionist approach is the need for the researcher(s) to acknowledge and examine their own involvement (including their beliefs and assumptions) in the research process. This in turn requires reflexivity on the part of the researcher(s) in order to understand, at least in part, the nature and significance of their influence on the research process and findings. The research team consisted of researchers from different cultural/ethnic backgrounds and some had a religious background whilst others did not. A range of different experiences and perspectives informed the design and implementation of the study.

The first author (MT) is a first-generation Vietnamese Australian female researcher with no religious background, however she has had some personal exposure to Buddhism within her family and Christianity during 2 years of attending a Christian high school. She conducted all interviews and focus groups in this study with the exception of one focus group that was facilitated by a co-author who is a female with an Afghani and Muslim background. Both researchers disclosed their position as researchers from a university, but did not explicitly mention their cultural background or other personal and professional details unless asked by the participants.

## Findings

Six main themes were identified describing attitudes, beliefs and knowledge about FDV within culturally diverse Australian faith communities: 1) Faith and religion do not condone violence; 2) Awareness of FDV is increasing, yet remains often poorly understood; 3) FDV is still a taboo topic; 4) Denial and defensiveness about FDV persist; 5) Patience, endurance and forgiveness is often prioritised over safety; 6) Gender roles and norms founded on religious beliefs and interpretations underpin many FDV understandings and responses.

As a result of the overlapping nature of some participants’ roles and experiences, the viewpoints of FDV sector workers, faith leaders and faith community members are integrated. For example, some workers are also members of a faith-based community, and some community members had previously worked in, or currently work in, the FDV sector. To protect the identity of participants, codes are used to identify the participant group i.e. worker (W), faith leader (FL) or community member (CM or CM_FG for focus group). Participants also have their faith-background noted.

### Faith and religion do not condone violence

It was strongly emphasised by faith leader participants that their faith and religion did not condone FDV, and that violence was abhorrent to their fundamental religious beliefs. They all saw FDV as associated with a misinterpretation of their beliefs, not inherent to their faith.

> *In Judaism its value system would be one that would completely abhor abusive relationships but nevertheless not everyone is in sync with those values*. FL9 (Jewish)
>
> *In Buddhist aspect we – in daily life we already teach them every day when you come you have to focus on the precept of Buddhism, it’s – if everyone have [sic] practice or precept, five precept of Buddhism, it’s no family violence*. FL10 (Buddhist)

A misinterpretation of faith was explained by a Christian faith leader in this way:

> *I think theologically not many people have put their time and energy into thinking about how some of the real basics of the Christian faith, the idea that Jesus died for us so that we would be forgiven by God and that because we’ve been forgiven by God we have the resources to forgive others. Some of those things I don’t think have kind of been passed through the domestic violence lens to say how do we teach that in a way that’s sensitive to how that is heard and kind of misused by abusers?* FL2 (Christian)

Some Muslim community members concurred with this view.

> *No, I think most of them [faith leaders] are you know in line with that it’s [violence] not allowed … that’s what Islam says, that this not condoned at all in our community*.CM_FG5(Muslim)
>
> *There are real cases in everywhere in the world, everywhere in the world, it’s not only the Muslim or Islamic world so it’s everywhere. It doesn’t have to [do] anything with the religions*. CM_FG1 (Muslim)

Several faith leaders and community members suggested that FDV was related to ethnic cultural practices rather than being directly related to faith and religion. They clearly distinguished ethnic culture as separate to faith and religion, especially for recent migrants to Australia, although the degree to which religion and culture were seen as separate or overlapping varied.

> *Because that’s their culture and it’s not their religion. Religion does not sanction this [violence] but certain culture, they bring from the village*. WI7 (and Sikh community member)
>
> *It’s the cultural nuances that have been added to the faith*. WI6
>
> *In terms of physical abuse, I know of a couple… she was a pastor’s wife and you know they come from a very strong Greek culture and so there was cultural stuff as well as spiritual stuff and the husband who was a pastor … would even physically hurt her as well as emotionally manipulate things and spiritually twist things from scripture so she never found that there was a right answer. ‘Cause I think there was cultural stuff and not just the spiritual stuff and the cultural stuff that was oppressing her, that was telling her to be a good Greek wife*… CM4 (Christian)

### Awareness of FDV is increasing, yet remains often poorly understood

Faith leaders and community members from all faiths recognised that FDV was an important issue both within their faith contexts as well as amongst the broader community.

> *I’d say it’s in [sic] – very prevalent for a good number of the women that I meet with just for pastoral or for training purposes end up disclosing abuse to me at some point in our relationship so it would be more common than not*. FL1 (Christian)
>
> *I think family violence is as prevalent within the Jewish community as it is in all other communities*. FL8 (Jewish)

Some community member participants from Muslim and Hindu faiths and some Jewish and Christian faith leaders, felt that awareness was increasing in their communities.

> *Now people have started talking, now that there is lot of awareness going on and especially the education with – among women when we talk about our community, now women have started knowing that this is not okay. If someone is abusing you it’s not okay*. CM_FG4 (Muslim & Hindu)
>
> *There’s been a very big push in the last number of years to raise awareness in the Jewish community on domestic violence and abusive relationships on all levels and so on, and that’s been very, very helpful*. FL9 (Jewish)
>
> *[I] think in the last 10 years I can witness that there’s a growing trend of openness, but still compared to the mainstream it’s still, we are quite far away*. FL3 (Christian)

Despite awareness increasing, leader and community member participants described that some in their community still had a very low knowledge and understanding of FDV. This included faith community members being unaware that they were experiencing FDV

> *Sometimes victims don’t even know that they’re being abused and that’s the thing*. CM_FG2 (Muslim and Christian)

A key knowledge gap identified was that understandings of FDV was often limited to extreme physical violence, with other forms of abuse, such as emotional, social, financial and spiritual abuse not recognized.

> *I don’t think the very wider perception about domestic violence were understood, but it’s more like the physical one, if my spouse is very violent, he’s always very angry, he throws stuff around, they understand. But in terms of issues like verbal violence or mental violence I don’t think so*. FL3 (Christian)

### FDV is still a taboo topic

Despite the increased awareness of the issue of FDV, participants commonly described FDV as a taboo topic within faith communities. This taboo was commonly attributed to views that family relationships were private, personal issues and shameful or inappropriate to discuss publicly.

> *I think it’s quite prevalent but it’s hidden… People just don’t talk about it, they try to keep it within the confines of their own four walls and only in extreme cases do they ask for help from outside. That’s my belief*. CM_FG2 (Muslim & Hindu FG, Muslim participant)
>
> *As Asian we too scared to talk. We keep it [to] ourselves. If we do have family problems then we just ask the monks … to do prayer for the family’s at peace (sic)*. CM1 (Buddhist)
>
> *I think there will definitely be still people in our church that still come from that place and probably not readily freely want to talk about these issues at all. I mean no-one likes to air their dirty laundry*. CM4 (Christian)

Some suggested that reluctance to discuss FDV is potentially greater in more conservative communities, again related to higher pressure to maintain community norms and not raise difficult issues publicly.

> *I think in a more secular Jewish context it’d be more likely to be spoken about than in a - ultra-religious one because you know it’s really the pressure to conform, the pressure to not rock the boat would be a lot higher, I would hazard a guess*. CM7 (Jewish)

### Denial and defensiveness about family and domestic violence persist

Faith leader and community member participants from Christian, Jewish and Muslim faiths discussed that denial and defensive attitudes about FDV remained high in their communities.

> *I think there are people who – that – whose church just deny that it was an issue at all and so didn’t have any training or staff available for people who’d experienced it*. FL2 (Christian)
>
> *Sometimes we underestimate the level of denial in these types of communities. It’s so strong*. CM_FG3 (Jewish)

Several participants related this denial and defensiveness to a perception that FDV would not be an issue if one is a true follower of faith.

> *The way that churches they … deal with it is you just need to get your theology right, and you need to be a loving Christian and abuse won’t be a problem in your life*. WI3
>
> *It’s other people who are in abusive relationships, not Christians*. WI1
>
> *Re other people’s attitudes: No, DV’s not an issue, good Muslim people don’t do that*. CM_FG2 (Muslim and Hindu FG, Muslim participant)

Denial, defensiveness and lack of acknowledgment of FDV as an issue among those with strong faith and religious practices were also identified by participants as resulting in victim-blaming attitudes and failing to believe women who disclose FDV experiences within faith contexts.

> *So then she drums up the courage and goes to her imam and the imam looks at her and ‘oh but he’s a very good Muslim and he comes to my congregation and he’s a good man and he’s this and he wouldn’t do that to you’*. WI4

### Patience, endurance and forgiveness is often prioritized over safety

Across many faiths, religious beliefs and interpretations of sacred texts that prioritize acceptance, endurance, forgiveness and maintenance of relationships over safety in situations of FDV was identified. These beliefs and interpretations were seen as contributing to shaming of those speaking out against FDV either personally or as community advocates, and to victim-blaming attitudes that placed responsibility for FDV on victims rather than perpetrators.

> *So a woman particularly has been taught from fairly young age if something bad happens to you, you have to accept it, you’ve got to forgive – say for example if you’re married in the context of family violence, the abuser was your husband or other members of the family. And if you don’t it means that you’re not strong enough so instead of – they link that strength in terms of forgiveness*. CM8 (Muslim)
>
> *Or blaming her or kind of saying ‘you know it takes two to tango’ and so ‘yes, he’s done wrong things but so have you and you need to repent of those and you need to apologize to him or you need to maintain contact with him or be open to reconciliation with him. You should be willing to forgive him’*. FL2 (Christian)
>
> *What we have is the Dharma, or what we call Buddhist teaching, and what we could do is to advise them to step back and look upon this from a karmic situation that certainly there could be something in this, in their previous life … we offer a space that they can come to, be away from abusive relationships and that the other party knows they’re in a safe space and give them time to reconcile*. FL7 (Buddhist)

### Gender roles and norms founded on religious beliefs and interpretations underpin many FDV understandings and responses

Beliefs and interpretations of sacred texts that prescribe and proscribe specific gender roles and norms were identified as core underpinnings of understandings and responses to FDV in many faith communities. Some participants described beliefs that strongly endorsed an ‘equal but different’ view of women and men, in which women and men were seen to have equal rights and value, but different roles and expectations for behaviour both within faith contexts and in wider society. In particular, interpretations that uphold men as active, leaders, and to inherently possess authority domestically and in public life, with women as more passive and submissive to men. How these different gender roles and expectations translated to equal rights and access to opportunities in practice was not, however, articulated.

> *So you know in Judaism … ritualistic – in terms of ritual men have a more active role in that area, women’s role is more passive in that area and some would interpret that as being sort of sexist or chauvinist or what have you, but when you actually speak to practitioners of the faith and religious people they’ll say that you know the women in their families and the women in their communities are valued equally but simply have a different kind of role*. FL8 (Jewish)
>
> *Even though if sometimes people would approach religious leader some of them might hold the view again that the husband is superior to the wife and the wife should just compromise*. CM6 (Hindu)
>
> *The language around kingship and around headship and around the pronouns that they use. And then it’s even sort of notions of them. As I said before about the unconscious assumptions about adulthood and males. I mean when you think of the way that language… and other norms normalise inequality*. WI1 (also Christian background)

Muslim and Christian participants identified that victim blaming, minimising and failing to believe FDV disclosures from women are even more an issue when the perpetrator has a high status within the community.

> *So this is when they realised that this [perpetrator] is a distinguished person in the community, this is someone who can help them in the hospital, this is someone who is qualified and you know qualified surgeon and all that so he’s someone they can benefit from so the family and everyone will actually have a soft corner for that because he will be of some advantage at some time in their life*. CM5 (Muslim)
>
> *[There were] instances where the abuser was a significant member of the church so even a minister or an elder in the church and … when the wife or partner chose to leave, spinning stories that made her seem like the person who’d given up on the relationship and was sort of unreasonably unwilling to consider reconciliation*. FL2 (Christian)

The nature of leadership structures and gender role norms within many faith contexts means that it is far more common for men to be in such high-status positions within faith and religious organizational contexts, as well as within the wider community.

## Discussion

This study furthers our understanding of the attitudes, beliefs and knowledge related to FDV within culturally diverse faith communities in Australia. Study findings demonstrate the tensions that exist between expressions of faith and attitudes to women and FDV. In particular, the general agreement among participants that their respective faiths did not condone violence while simultaneously identifying cultural structures within their faiths that enabled and ignored abuse against women. While many participants described continued denial and defensiveness about FDV in their communities where the topic remains taboo, many also felt that awareness of FDV was increasing.

Many findings of this Australian study are consistent with those of previous work internationally, and reinforce the complex relationships between religion and faith and FDV as well as with migration and culture. A strong belief that violence is not condoned by religion and that FDV is a doctrinal misunderstanding has been identified as a common view (Aune & Barnes, 2018; Haaken, Fussell, & Mankowski, 2007; Ibrahim, 2020). The extent to which faith leaders and communities can address FDV without engaging with these structural and systemic issues has been questioned and remains a key outstanding issue (Holmes, 2012; Murdolo & Quiazon, 2016; Priest, 2018; Vaughan et al., 2020a; Westenberg, 2017).

Participants in this study identified that awareness of FDV was increasing in their faith communities, although overall awareness remained low. The need for a more expansive understanding of FDV beyond physical violence to include social, emotional, financial and spiritual abuse was identified. This is consistent with a recent report from the US (Sojourners & IMA World Health, June 2014) that showed awareness of FDV is increasing among Christian pastors. A small UK study among churches also found views that framed FDV as physical violence and a lack of understanding of emotional abuse (Aune & Barnes, 2018). Both of these studies identified a need for training faith leaders in identifying and responding to FDV, echoing earlier calls for training of faith leaders, including in seminary programs (Drumm et al., 2018; Jones et al., 2006; McMullin, 2018).

Despite growing awareness, denial and defensiveness about the presence of FDV in faith communities was reported. For example, the view that faith and FDV are antithetical has been identified by survivors as a key form of denial and defensiveness about the presence of FDV faith communities (Aune & Barnes, 2018). Views that frame FDV as a doctrinal misunderstanding or individual sin rather than as a systemic, structural issue continue to contribute to denial and defensiveness. Empirical work from the UK as well as Australia suggests that faith community members recognise that FDV is an issue in wider society but are far less likely to consider it an issue in their own communities (Aune & Barnes, 2018; Perry & Priest, In preparation).

At the institutional level, the view that family and marriage relationships are private issues that should not be discussed outside of the home may create barriers to help-seeking for those experiencing violence. This is consistent with a study among US African Christian migrants that found prioritisation of marriage and community reputation, avoidance of gossip or community shame, and a view that marital issues were private matters all contributed to low levels of help seeking for FDV among these women (Ting & Panchanadeswaran, 2016). Similarly, a study among Adventist women in the US found that beliefs around marriage and divorce can create multiple barriers to help-seeking for FDV (Popescu et al., 2009). Others have also emphasised that beliefs in the sanctity of marriage along with anti-divorce values can act as barriers in FDV help-seeking (Hassouneh-Phillips, 2001; Nash, Faulkner, & Abell, 2013).

Another institutional issue identified was the prioritisation of forgiveness, endurance and patience over safety. Previous studies have similarly identified that faith leaders can encourage those experiencing abuse to foster forgiveness and endurance and to stay in abusive relationships and/or fail to refer those experiencing FDV to support services and that this is supported by theological and doctrinal beliefs and language (Ghafournia, 2017; Ringel & Park, 2008; Westenberg, 2017). It has also been identified elsewhere that faith leaders advise reconciliation or marriage counselling in situations of abuse with low awareness that doing so is inappropriate and may endanger a woman’s safety (Choi, 2015; Miles, 2000).

A major theme in this study, and in the literature more broadly, is how patriarchal culture and religious norms about gender roles contribute to FDV in faith communities. This was reinforced in this study across Christian, Jewish and Muslim communities in particular. Previous work has identified that gender norms and doctrinal beliefs that encourage women to be subservient and men to be dominant are important factors impacting the extent to which women in faith communities recognise experiences of FDV, seek help for FDV, and to which they are supported appropriately (Fortune, Abugideiri, & Dratch, 2010; Ringel & Bina, 2007; Ting & Panchanadeswaran, 2016; Wendt, 2009). Some participants in this study identified the ‘equal but different’ roles for men and women described in some faith cultures as a significant issue. Facilitating interpretations of male leadership and female submission, doctrinal beliefs in God-ordained gender hierarchical roles may contribute to and perpetuate FDV at interpersonal and institutional levels (Haaken et al., 2007; Westenberg, 2017). Exploring how to sensitively and effectively increase recognition within faith communities of the ways in which gender inequalities underlay FDV at individual and institutional levels is an important area for future work (Webster & Flood, 2015). In particular, messages addressing gender equality issues need to be tailored to avoid conflict between mainstream FDV frameworks and religious teachings specific to each faith community and context (Le Roux & Loots, 2017; Westenberg, 2017).

### Limitations

There are several limitations to this study. Firstly, the use of qualitative methods means the results are not intended to be generalisable to any particular group or community based on their cultural or religious background. Whilst a broad range of experiences and perspectives was captured across different religious and cultural backgrounds, it is possible that some points of view were not included as well as that some were overstated. Secondly, the majority of community member interviews and focus groups were with women, therefore, the range of views held by different men in the community were not fully captured and highlights an area for future research. Finally, this work was limited to urban areas across four Australian states. The different communities, service systems, policies, and immigration histories from other regions and states may provide slightly different or additional experiences that were not captured in this project.

Further exploration of all of the aforementioned issues within specific faith communities, as well as how to support and engage with these communities in increasing understandings of FDV and developing effective responses, is needed in an Australian context.

## Data Availability

n/a

